# Neural and behavioural measures from attention testing show no support for efficacy of neurofeedback treatment for adult ADHD

**DOI:** 10.64898/2026.04.26.26351764

**Authors:** Jinyu Wang, Andrei E D Rodionov, Benjamin Ultan Cowley

## Abstract

Attention-deficit/hyperactivity disorder (ADHD) is associated with impairments in sustained attention and inhibitory control. Neurofeedback (NFB) is a widely used non-pharmacological treatment for ADHD and is generally well tolerated, but evidence for its efficacy remains mixed. Here we report results from secondary analysis of a randomized controlled trial of NFB training for adult ADHD, analysing behaviour and neural data from attention testing in both test-retest and treatment-vs-waiting list control group contrasts. We used electroencephalography (EEG) to investigate event-related cortical dynamics during the Test of Variables of Attention (TOVA), administered before and after NFB treatment. 44 adults with ADHD (NFB treatment, ADHD-T: n = 23; waitlist control, ADHD-W: n = 21) completed the TOVA before and after the NFB training period, while 128-channel EEG was recorded. Treatment-related change was examined through analyses based on behavioural TOVA performance, power spectral density, and event-related potentials, analysed with Bayesian linear mixed models. We found no meaningful evidence for NFB-specific improvements in TOVA behavioural performance over time, and no evidence that NFB modulated ERP or spectral indices relative to the ADHD-W group. Overall, we found no evidence that NFB treatment meaningfully benefited sustained attention or inhibitory control in adults with ADHD.

## Introduction

Attention Deficit/Hyperactivity Disorder (ADHD) is a common psychiatric disorder characterized by inattention, hyperactivity, and/or impulsive behaviour (Volkow & Swanson, 2013). Although typically observed in childhood, ADHD affects 4-5% of adults (Biederman, 2005; Kessler et al., 2005), and about 65% of children diagnosed with ADHD later experience symptoms in adulthood (Faraone et al., 2006). EEG-neurofeedback (NFB) is a widely-practised non-pharmacological training treatment for adults with ADHD. NFB has been applied to treat ADHD for over 50 years (Lubar & Shouse, 1976), and is based on a brain-computer interface that processes real-time EEG signals, targeting specific frequency bands or brain potentials for self-regulation (Enriquez-Geppert et al., 2017). However, in spite of many randomized controlled trials (RCTs), the evidence for NFB efficacy is mixed and inconsistent (Kuznetsova et al., 2023). Initial meta-analyses suggested a moderate to strong effect size for NFB efficacy on ADHD symptomatology (Arns et al., 2009; Hodgson et al., 2014), but this was challenged when probably-blinded outcomes were analysed (Bussalb et al., 2019; Cortese et al., 2016). Most recently, a meta-analysis of 38 RCTs with probably-blinded outcomes found no significant improvement in ADHD symptoms (Westwood et al., 2025).

Null RCT results are, however, contradicted by considerable anecdotal clinical evidence for the efficacy of NFB – in other words, NFB treatment ‘in the wild’ very often seems to work. Thus, the case for NFB efficacy remains unclear and the number of well-controlled RCTs (cf. Arnold et al., 2021) suggest that the lack of understanding lies with the specific mechanisms of effect linking treatment to ADHD symptoms. Put another way: standard RCT designs, while effective for pharmacological trials, may be limited in capturing the therapeutic impact of NFB. Prior NFB RCTs have aimed to isolate feedback as a pure form of neural operant conditioning. However, the efficacy of NFB may also depend on the *interaction* between contingent feedback and various controlled factors such as trainer interaction (Kuznetsova et al., 2026), which could lead to an underestimation or obfuscation of treatment effects in well-controlled designs. A complementary approach is thus to evaluate whether NFB produces changes in the neurocognitive domains most affected by ADHD, such as sustained attention, using pre-post assessments of behavioural performance and their neural correlates. Here, we report such a study, to determine if NFB engages specific neurocognitive mechanisms regardless of changes in global symptom ratings.

Attentional fluctuation in ADHD is commonly reflected in reduced task-evoked neurophysiological measures of attention such as attention-related ERPs. These neurophysiological measures reliably differentiate ADHD from healthy controls, with reduced ERP amplitudes (N2, P3) and altered spectral patterns reflecting core deficits in attentional control and inhibitory processes (Hasler et al., 2016; Loo & Makeig, 2012). Moreover, these neural correlates are task dependent, shifting with task demands and repeated exposure (Cheung et al., 2017), making them reliable mechanistic readouts for examining *whether non-pharmacological treatments engage and change the targeted neurocognitive processes*. While early studies reported neurophysiological changes following NFB, such as increased contingent negative variation (CNV) in children with ADHD (Wangler et al., 2011) and reduced reaction time variability (RTV) in adults with ADHD (Mayer et al., 2016), subsequent studies reported mixed or null findings. Deiber et al. (2021) found increased N1 and P3 amplitudes after training, but these changes occurred equally in healthy controls. Similarly, Aggensteiner et al. (2021) found no treatment-specific changes in behavioural performance or ERPs (P3, CNV) during a CPT, when comparing NFB to a semi-active control condition. Thus, it remains unclear whether NFB produces specific changes to attention-related neurocognitive processes in ADHD.

Here, we investigate behavioural performance and task-evoked neurophysiological responses during a sustained-attention task given before and after 40 sessions of ∼1h-long NFB treatment. Data came from an RCT that investigated NFB treatment for adult ADHD (Cowley et al., 2016), where training regimes included the classic protocols theta-beta ratio (TBR) and sensorimotor rhythm (SMR), as well as inverse-target versions of these^1^. The RCT had two measurement time points: *intake* (baseline measurements taken before treatment), involving the initially recruited ADHD group and a matched neurotypical control (NTC) group; and *outtake* (measurements after treatment), wherein the ADHD group had been divided into a treatment group and a waiting list control group. Both measurement times included EEG recordings during the Test of Variables of Attention (TOVA, Greenberg et al., 2016).

CPTs have been widely used as assessment tools in NFB intervention for neurotypical (Egner & Gruzelier, 2004) and ADHD population (Alegria et al., 2017; Perreau-Linck et al., 2010; Schneider et al., 2022; Schönenberg et al., 2017). As a well-established CPT, TOVA is designed to measure sustained attention. Based on a normative database of 1596 individuals, TOVA tests the speed and accuracy of attention processing. Recognized as the gold-standard behavioural paradigm for attention deficit assessment, it has been used to investigate sustained attention in children and adolescents diagnosed with ADHD (Medici et al., 2018; O’Connell et al., 2009; Wu et al., 2007). However, fewer studies have focused on ADHD adults (Grane et al., 2014; Memória et al., 2018), and measuring neural activities during TOVA in ADHD has been done only twice, to our knowledge (Cowley et al., 2022; Halawa et al., 2017).

Here, we address the core question, *what is the long-term impact of NFB treatment on neural correlates of sustained attention in adult ADHD?* We examine test-retest results (intake–outtake) on behavioural and neurophysiological differences between the ADHD-T and ADHD-W groups. We test TOVA behavioural performance, *α* and *θ* power spectral density (PSD), and cognitive ERP components (N2/P3). If NFB produces specific treatment effects, we would expect the ADHD-T group to show differential improvements (relative to the ADHD-W group) in these measures of attention processing. Our results do not support that expectation.

## Results

Participants completed TOVA as the first CPT in their EEG recording session. TOVA takes 22.6 minutes, and while originally designed as a behavioural attention test, not a brain imaging protocol, its characteristics as a Go/Nogo target-classification task suggest that it can be suited for assessing atypical event-related power in ADHD (Michelini, Salmastyan, et al., 2022). Go and Nogo trials are distributed in a ratio of 2:7 which inverts halfway through the test. Thus, with infrequent Go trials, H1 tests vigilance (challenges inattention), while with frequent Go trials, H2 tests inhibition (challenges hyperactivity).

Data analysed below include those ADHD participants (N = 44) measured at intake *and* outtake; also the matched NTC group, measured once (N = 18, 12 females, 6 males). From an initial sample of N = 54 (29 females, 25 males), the ADHD group had been randomised into a treatment group (ADHD-T, n = 23, 2 dropouts from intake) and a waiting list control group (ADHD-W, n = 21, 8 dropouts), according to constrained randomisation procedure that balanced age and gender.

### Behavioural Performance

TOVA returns five standardised variables (see Figure 1 for distributions; see Materials and Methods for derivation): mean response time (MRT), response time variability (RTV), commission errors (COM), omission errors (OM); and *d′*^2^. To evaluate whether NFB treatment improved TOVA performance, we modelled the five TOVA variables using Bayesian LMMs (see detailed descriptive statistics in Supplementary Table S1). The primary test of NFB efficacy was the Group × Time interaction (*difference-in-differences*). Posterior estimates for Group × Time were small and uncertain across measures (95% Crls spanning zero, see Table 1), *indicating little evidence for NFB-specific improvement*^3^.

**Figure 1:**
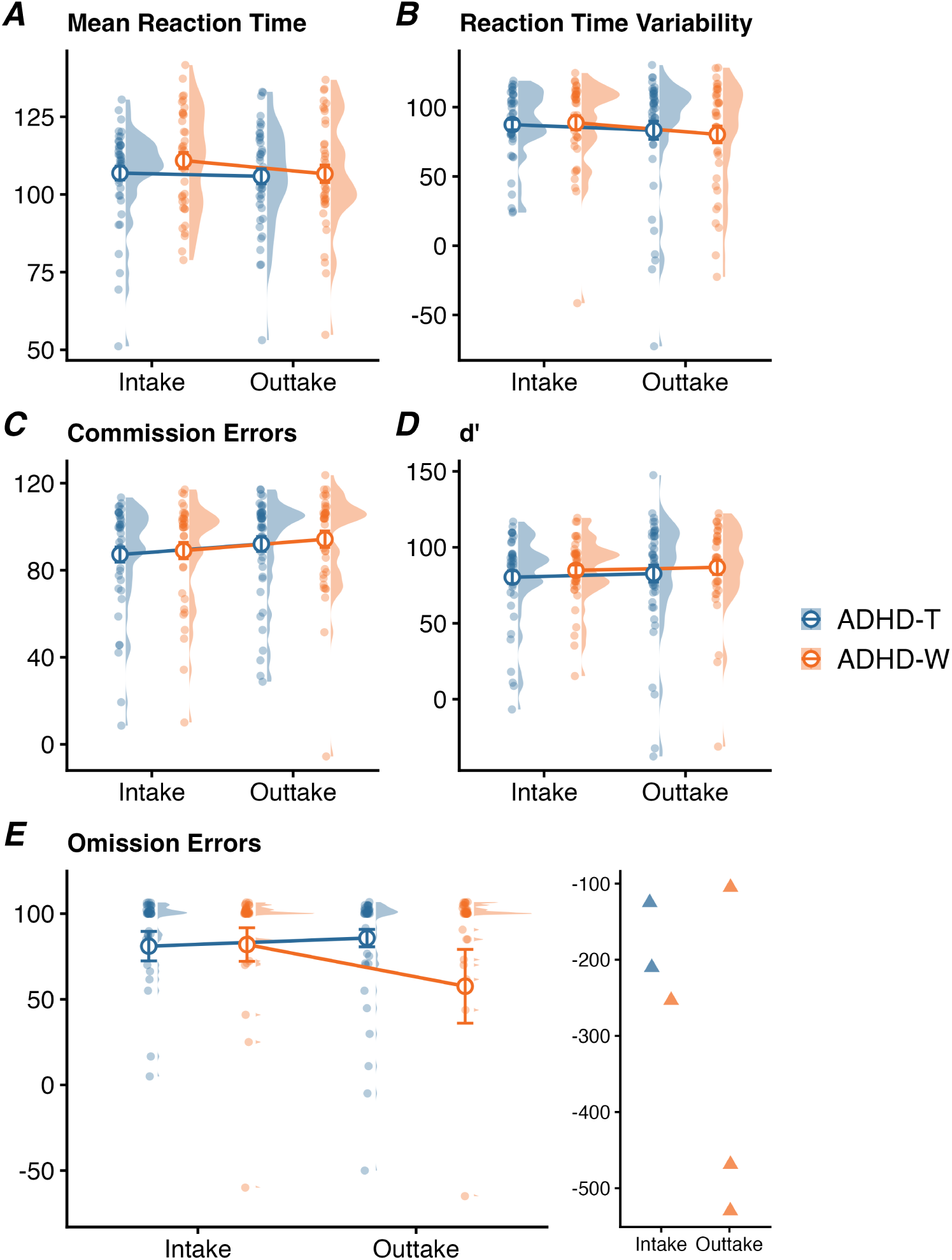
Behavioural performance on the TOVA task. All measures are TOVA standard scores, where values *<*100 indicate worse performance than the normative database. Raincloud plots show distributions for ADHD-T (blue) and ADHD-W (orange) at intake and outtake. **(A)** mean reaction time, **(B)** reaction time variability, **(C)** commission errors, **(D)** d-prime (*d^′^*), **(E)** omission errors. The worse performance in omission errors (n = 6, 3.5%) are displayed separately in the right inset due to y-axis constraints; these values were retained in statistical analyses. **Note 1**: Nine additional outlier scores, identified via studentized residuals, were excluded across all five variables prior to analysis (see Materials and Methods). Large circles represent group means with standard error bars. **Note 2**: This figure displays descriptive statistics; Bayesian LMM results are reported in the main text and Table 1.

**Table 1:** Bayesian linear mixed model results for TOVA performance variables.

| Effect | Dependent Variables |  |  |  |  |
| --- | --- | --- | --- | --- | --- |
| | MRT | RTV | COM | OM | $d'$ |
| Group (ADHD-W vs. ADHD-T) | 0.19<br>[−0.19, 0.56] | 0.03<br>[−0.15, 0.20] | 0.20<br>[−0.13, 0.54] | −0.02<br>[−0.08, 0.03] | 0.15<br>[−0.13, 0.42] |
| Time (*Outtake* vs. *Intake*) | 0.00<br>[−0.29, 0.28] | −0.06<br>[−0.25, 0.13] | 0.04<br>[−0.27, 0.35] | −0.03<br>[−0.09, 0.03] | 0.24<br>[0.01, 0.47] |
| Condition (H2 vs. H1) | 0.02<br>[−0.15, 0.20] | −0.01<br>[−0.19, 0.16] | −0.15<br>[−0.46, 0.13] | −0.04<br>[−0.11, 0.02] | −0.08<br>[−0.32, 0.14] |
| Group × Time | −0.22<br>[−0.63, 0.19] | −0.22<br>[−0.49, 0.05] | −0.04<br>[−0.46, 0.40] | 0.02<br>[−0.06, 0.11] | −0.05<br>[−0.38, 0.27] |
| Group × Time × Condition | 0.33<br>[−0.03, 0.69] | 0.18<br>[−0.18, 0.53] | **0.74**<br>**[0.15, 1.34]** | 0.00<br>[−0.12, 0.13] | 0.26<br>[−0.18, 0.69] |

*Note.* MRT = mean response time; RTV = reaction time variability; COM = commission errors; OM = omission errors; $d'$ = d-prime. Values are posterior estimates with 95% credible intervals in brackets. Reference: Group = ADHD-T; Time = intake; TOVA Condition = H1 (infrequent targets). Models controlled for API.

The only interaction whose 95% Crl excluded zero was a Group × Time × TOVA condition effect on commission errors (*β* = 0.74, 95% Crl [0.15, 1.34]). Follow-up estimated marginal means showed that this interaction was specific to TOVA second half (H2), where the standard scores of commission errors increased (indicating fewer errors) from intake to outtake in the ADHD-W group (Δ_outtake_*_−_*_intake_ = 0.51, 95% Crl [0.17, 0.86]), whereas change in the ADHD-T group was uncertain (Δ_outtake_*_−_*_intake_ = −0.19, 95% Crl [−0.57, 0.17]). Consistently, the between-group difference in change (difference-in-differences) was supported in H2 (ΔΔ = 0.71, 95% Crl [0.20, 1.22]) but not in the first half (H1) (ΔΔ = −0.04, 95% Crl [−0.47, 0.39]). This pattern showed a relative improved performance in the ADHD-W group rather than clear improvement in the ADHD-T group. In addition, *d^′^* increased modestly over time across both groups (Time: *β*, = 0.24 95% CrI [0.01, 0.47]) with little evidence for a group difference in change (Group × Time: *β* = −0.05, 95% Crl [−0.38, 0.27]).

### ERP analysis

We analysed 2000 ms EEG epochs, time-locked 1000 ms before and after the target stimulus onset, and baseline corrected. Epoch sets were divided between TOVA conditions: correct responses and correct inhibitions, and TOVA halves H1 and H2, to give four datasets per group for most analyses.

Across groups, TOVA condition, and frontal and parietal regions of interest (ROIs), N2 amplitudes ranged from −0.90 to 0.16 *µ*V and P3 amplitudes ranged from 1.24 to 5.13 *µ*V. Peak latencies remained relatively stable, centring approximately at 200–220 ms for the N2 and 370–400 ms for the P3. Visual inspection of grand-average ERPs waveforms in both ADHD groups across intake and outtake is shown in Figure 2A. Although visual inspection shows a general increase in P3 amplitude from intake to outtake in ADHD-T group, a comparable increase was also evident in the ADHD-W group. To examine potential treatment effects, we fitted four Bayesian LMMs to N2 and P3 mean amplitudes and peak latencies in target-locked correct-response trials. Across all four outcome measures, Group × Time interaction estimates were close to zero and their 95% Crls included 0 (N2 mean amplitude: *β*_Group_*_×_*_Time_ = −0.01 *µ*V, 95% Crl [−0.42, 0.41]; N2 peak latency: *β* = −1.41 ms, 95% Crl [−15.78, 12.87]; P3 mean amplitude: *β* = −0.01 *µ*V, 95% Crl [−0.47, 0.45]; P3 peak latency: *β* = 0.09 ms, 95% Crl [−11.80, 11.89]; Table S1). Posterior estimates for these interaction effects are shown in Figure 2C.

**Figure 2:**
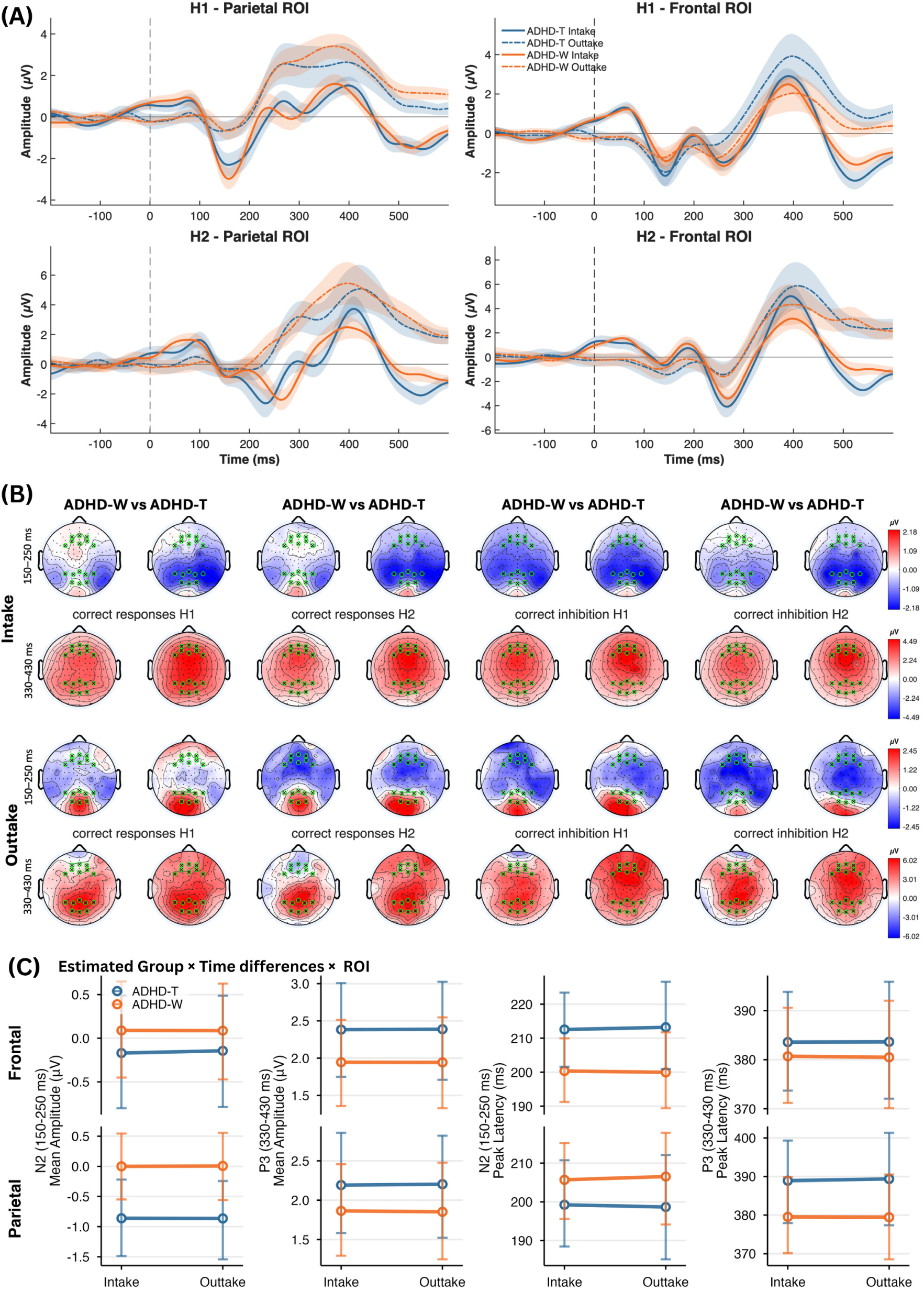
ERP waveforms and Bayesian LMM estimates for target-locked correct-response trials. (**A**) Grand-average ERPs in parietal and frontal ROIs for H1 and H2 conditions. Solid lines denote intake and dashed lines denote outtake; shaded areas indicate SEM. (**B**) Whole-head scalp maps of voltage averaged over same time windows (150–250 ms, rows 1,3; 330–430 ms, rows 2,4) used for testing ERPs. Each condition-response vs. inhibition and H1 vs. H2 - is shown at intake (rows 1–2) and outtake (rows 3–4). Colour scales are locked across each time window. ROI electrodes are shown as green crossed circles. (**C**) Posterior mean estimates and 95% CrIs from Bayesian LMMs for N2 and P3 mean amplitudes and peak latencies in frontal and parietal ROIs Blue denotes ADHD-T and orange denotes ADHD-W.

Main effects of Group and Time were similarly negligible. For N2 mean amplitude, group differences (*β*_Group_ = 0.24 *µ*V, 95% Crl [−0.55, 1.02]) and time effects (*β*_Time_ = 0.01 *µ*V, 95% Crl [−0.33, 0.34]) showed no credible evidence of change. P3 mean amplitude showed comparable null effects (Group: *β* = −0.43 *µ*V, 95% Crl [−1.22, 0.38]; Time: *β* = 0.01 *µ*V, 95% CrI [−0.35, 0.36]). N2 and P3 peak latencies similarly showed no group or time effects (all 95% Crls included zero). Full posterior summaries for all fixed effects are reported in Supplementary Materials (Table S3). Additionally, because the primary ERP models showed a consistent main effect of TOVA condition on P3 mean amplitude and peak latency (see Table S3), we ran sensitivity analyses in which TOVA half (H1 vs. H2) was allowed to moderate the Group × Time effect. We fitted two extended models with Group × Time × TOVA condition interaction for mean amplitude and latency. Across both models, the three-way interaction estimates were small and their 95% Crls all included zero. TOVA condition did not evidence any moderating effect.

Visual inspection of the scalp topographies confirmed N2-like posterior negativity and P3-like centro-/fronto-parietal positivity in all conditions (Figure 2B). At intake, the ADHD-T group appeared to show stronger posterior negativity in the N2 window and more pronounced fronto-parietal positivity in the P3 window during correct response trials than the ADHD-W group. However, these group differences did not follow a consistent pattern when comparing intake with outtake, task conditions (response vs. inhibition), or TOVA halves. At outtake, no consistent spatial pattern emerged that reliably distinguished the two ADHD groups for any task half or response type.

### Spectral analysis

To test whether NFB modulated oscillatory activity, we analysed log-PSD at 4 Hz and 8 Hz during baseline and post-stimulus periods across ROIs and TOVA halves (Figure 3). We selected these frequencies based on their importance in prior work on the same task (Cowley et al., 2022). Across ROIs and TOVA conditions, log-PSD appeared slightly lower at outtake than intake for both baseline and post-stimulus periods; critically, Bayesian LMMs provided no evidence for a treatment-specific change. The four-way interaction indicated *β* = 0.16, 95% Crl [−1.22, 1.55] for 4 Hz, and *β* = −0.00, 95% Crl [−1.78, 1.78] for 8 Hz, with all Crls containing zero. Full posterior summaries for all fixed effects are reported in Supplementary Materials (Table S4).

**Figure 3:**
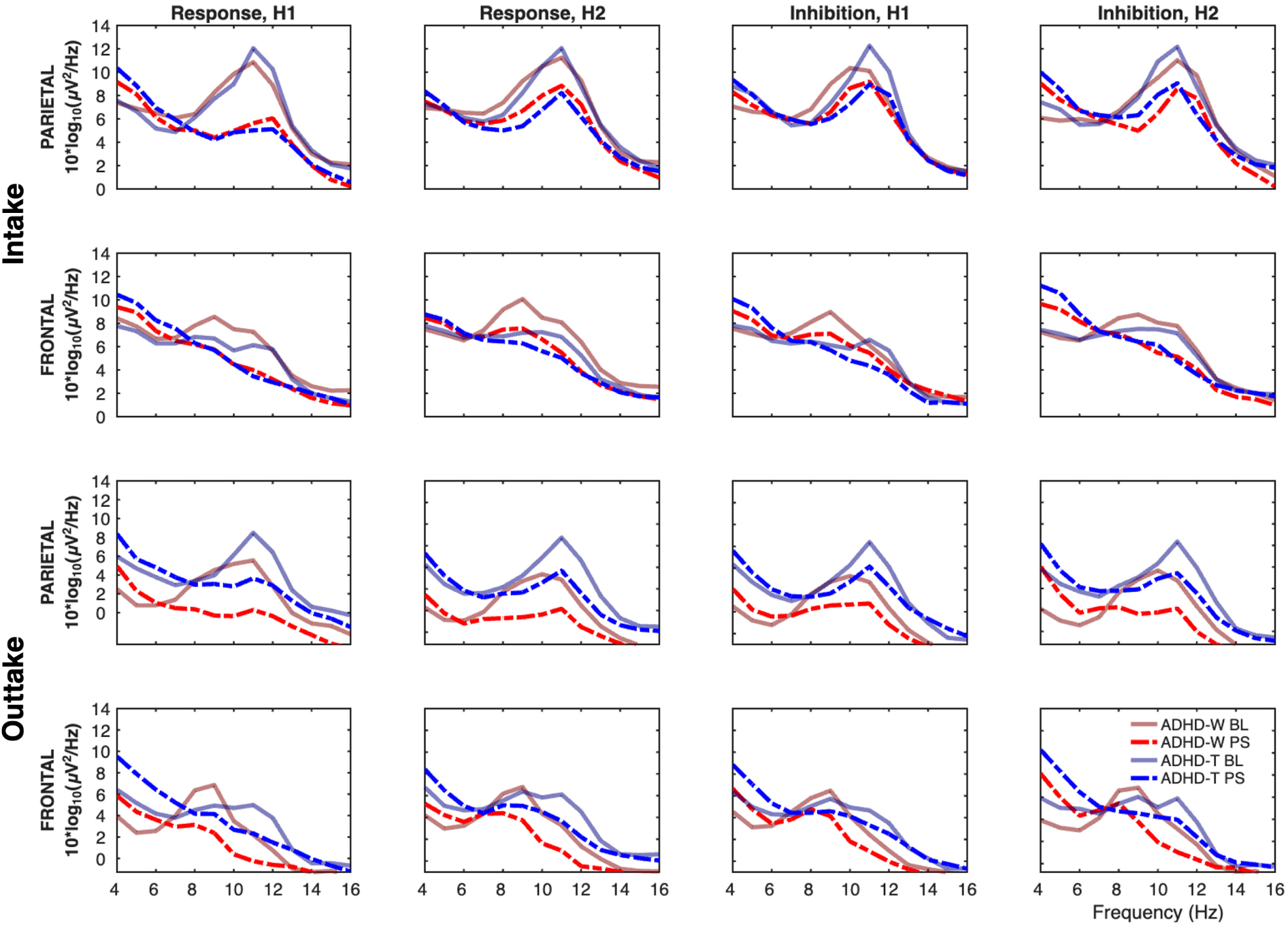
Log spectral power density (log-PSD) comparison between Intake (upper two rows) and Outtake (lower two rows). Log-PSD calculated during baseline (BL; brown and gray solid lines) and post-stimulus (PS; red and blue dashed lines) periods for ADHD-W (red) and ADHD-T (blue) groups, shown separately by ROIs (Parietal, rows 1,3 vs. Frontal, rows 2,4), Task Condition (Response, columns 1-2 vs. Inhibition, columns 3-4, and TOVA halves H1, columns 1,3 vs. H2, columns 2,4). The results indicate a prominent peak in the *α* band (∼10 Hz) during the Baseline period (solid lines), particularly in parietal region.

To further examine whether treatment modulated pre-stimulus *α*-band activity, we fitted another Bayesian LMM to parietal *α*-band power (8–12 Hz) on correct trials. The Group × Time interaction showed little evidence of NFB effects (*β* = −1.58, 95% Crl [−5.05, 1.89]). The results across individual frequencies in the *α* band confirm the null finding: 9 Hz (*β* = 0.17 [−0.93, 1.28]), 10 Hz (*β* = 0.01 [−1.11, 1.12]), 11 Hz (*β* = −0.55 [−1.66, 0.57]), 12 Hz (*β* = −1.08 [−2.19, 0.03]). NFB training did not differentially affect pre-stimulus *α*-band power between groups. These spectral results align with our behavioural and ERP findings, collectively demonstrating no NFB treatment efficacy.

### Comparison to neurotypical controls

In addition to contrasting ADHD-T and -W groups in their change before and after the NFB intervention, we contrast them *relative* to our NTC sample, in TOVA performance variables and ERP N2 and P3 mean amplitudes and latencies.

We fitted a Bayesian LMM comparing ADHD-T and ADHD-W intake and outtake against the NTC group for all five TOVA behavioural performance variables, separately for H1 and H2; see visualisation of results in Figure S1, Supplementary Materials. Posterior contrasts mainly showed a pattern of mild convergence between both ADHD groups and NTCs, with mostly either reduced or equal difference scores at outtake compared to intake. The only systematic exception was *d^′^*. Both groups showed lower standardized scores than NTCs during intake H1 (ADHD-T *β* = −0.381, 95% Crl [−0.670, −0.101]; ADHD-W *β* = −0.283, 95% Crl [−0.556, −0.004]), which converged at outtake H1; while both groups had equivalent *d^′^* to NTCs at intake H2, which improved non-credibly above NTCs at outtake H2. No other variable had systematic patterns of change from intake to outtake.

We fitted two Bayesian LMMs for ERP mean amplitude and peak latency at parietal and frontal ROIs, comparing the two ADHD groups at intake and outtake against the NTC group, separately for H1 and H2. The results are shown in Figure S2 in Supplementary Materials.

The posterior contrasts of mean amplitudes showed very similar patterns for both ADHD groups, during both TOVA halves, at frontal P3 and parietal N2: the difference from NTCs increased substantially from intake to outtake, as all outtake contrasts were credible (frontal P3: *β* from −2.39 to −2.85 *µ*V; parietal N2: *β* from 1.69 to 3.10 *µ*V). In contrast, the frontal N2 did not change much between times, and was only credibly different from NTC for ADHD-T at outtake in H1 (*β* = 1.290, 95% Crl [0.329, 2.231]). The one point of difference between groups in amplitude patterns was in parietal P3, where ADHD-T had minimal difference to NTC at intake in H1 (*β* = −0.131, 95% Crl [−1.137, 0.930])(likewise at both times in H2; intake: *β* = −0.617, 95% Crl [−1.702, 0.512], outtake: *β* = −0.919, 95% Crl [−1.926, 0.046]), but at outtake reached credible difference in H1 (*β* = −2.053, 95% Crl [−3.153, −0.941]); while ADHD-W difference to NTC went from negative at intake to positive at outtake, in both halves (H1: intake *β* = −0.523, 95% Crl [−1.559, 0.497], outtake *β* = 1.021, 95% Crl[-0.328, 2.268]; H2: intake *β* = −1.066, 95% Crl [−2.030, −0.114], outtake *β* = 0.713, 95% Crl [−0.391, 1.876]), credible only at intake H2.

For peak latency, differences relative to NTCs were generally small and non-credible, with only one contrast reaching the 95% Crl threshold: delayed parietal P3 latency during H1 in ADHD-W at intake (*β* = 24.436, 95% CrI [3.801, 46.836]). No consistent pattern emerged across ROIs, task halves, or timepoints.

## Discussion

The present study aimed to determine whether NFB modulates event-related neurocognitive activities underlying sustained attention in adults with ADHD. While we did not observe treatment-specific changes in either TOVA behavioural performance or event-related neural dynamics, we did observe modest non-specific changes over time: *d^′^* improved across both groups, and commission errors in H2 improved selectively in the ADHD-W group, both of which likely reflect practice effects rather than treatment efficacy.

### TOVA Performance

We re-analysed the behavioural TOVA performance using Bayesian LMMs to retest the null effects initially reported by Cowley et al. (2016), and extended this to also test each ADHD group against NTC performance.

We found that the TOVA did not reveal significant performance differences between the two ADHD groups before and after the treatment, or by contrast to NTCs. However, we found a credible Group × Time interaction for commission errors in the H2 condition. Specifically, this interaction was driven by a significant improvement in the ADHD-W group (standard score increase), whereas the ADHD-T group remained stable. Additionally, *d^′^* improved modestly over time across both groups, especially in H1 with respect to NTC. These behavioural changes likely reflect practice effects or increased task familiarity over the ∼8-month test interval, rather than NFB-specific efficacy. Our behavioural findings highlight the complexity of ADHD as a heterogeneous condition, where NFB treatment effects on sustained attention may vary substantially between individuals (Kuznetsova et al., 2023, 2026).

### Response Amplitude and Spectral Dynamics

Our analyses indicated no evidence that NFB modulated attention-related ERPs during TOVA performance. In the literature, reports of ERP modulation following NFB are mixed: effects vary with treatment protocols and task contexts and are not consistently treatment-specific under active or sham controls (Miranda et al., 2020). Visual inspection of the grand-average waveforms (Figure 2) suggested an overall increase in P3 amplitude at outtake in the ADHD-T group; however, similar retest-related changes were also evident in the ADHD-W group. In a further exploratory outtake-only comparison, the ADHD-T group showed larger frontal P3 amplitudes than the ADHD-W group in the target-locked condition (see Supplementary Materials, Figure S3). Nevertheless, the Bayesian analyses did not support any treatment-specific ERP modulation: both N2 and P3 remained unchanged relative to ADHD-W, with the null pattern consistent across mean amplitude and peak latency, frontal and parietal ROIs, and TOVA conditions (H1 and H2). Contrast with NTCs, showing the dynamics of change across groups, time, and halves with respect to a fixed reference point, also illustrated similar divergence from NTCs over time across ADHD groups and no evidence of a NFB treatment effect.

The above null pattern is compatible with boundary conditions on detectability in tasks that require sustained attention: although P3 and CNV are well-established indicators of attentional and inhibitory processing and are standard outcome measures in NFB research, they may provide a relatively insensitive readout of NFB treatment targets when measured during CPT demands (Varela et al., 2024), and may also shift with repeated testing, masking small effects under rigorous controls (Cheung et al., 2017). Accordingly, the modest improvement in ASRS self-report that we found after NFB treatment (reported in Cowley et al., 2016), may reflect compensatory strategy use or stabilisation of vigilance/arousal during daily functioning that does not necessarily transfer into detectable ERP changes during task performance.

In terms of the spectral power analysis, Bayesian LMMs revealed no evidence that NFB modulated oscillatory activity during TOVA. There were no differential treatment effects for either 4 Hz or 8 Hz power during baseline or post-stimulus periods; nor did pre-stimulus parietal *α*-band power show treatment-related changes. These spectral activity results are particularly notable given evidence linking *θ* oscillations to rhythmic attention sampling (Dugué et al., 2015; Haigh & Buckby, 2024). As proposed in Cowley et al. (2022), the periodic nature of TOVA task facilitates rhythmic sampling: a single target location is monitored at an 8 Hz rhythm, shifting to 4 Hz when attention must alternate between two locations (Holcombe & Chen, 2013; Landau & Fries, 2012). This sampling pattern is part of a more general modulation of *γ*-band activity (Landau et al., 2015) by *θ* oscillations through a process known as theta-gamma coupling (TGC) (Lisman & Jensen, 2013). TGC is a mechanism critical for memory encoding and sequential information retrieval, enabling the brain to represent and process multiple items in a defined order (Lisman & Buzsáki, 2008). TOVA sequentially involves monitoring stimulus spatial location by presenting two spatially distinct (but otherwise identical) stimuli in a distraction-free context, thus providing ideal conditions to generate strong ∼4-Hz parietal *θ* ERS, as empirically demonstrated at intake.

### Limitations and future studies

The trial was not double-blinded, which limits our ability to separate treatment-specific effects from non-specific influences such as expectancy. However, recent meta-analysis indicates that NFB effects do not vary as a function of how active the comparator arm is, with effects not observed even in trials with less active controls (Westwood et al., 2025). Additionally, recent work on the mechanisms of learning in NFB have suggested that treatment involves not just operant conditioning but also skill acquisition (Kuznetsova et al., 2026; Veilahti et al., 2021), and thus may actually *depend* on non-specific influences. This means that both active and non-active controls should be investigated to elucidate the neural effects of NFB, if any. Future studies extending our approach would complement our results and broaden general understanding by focusing on pre-post data from designs that feature both waiting-list *and* sham- or active-control designs, with blinded outcome assessment.

While statistical sensitivity, especially for neurophysiological outcomes, was affected by attrition and exclusions of low-quality data, resulting in smaller effective sample sizes for analyses, our null findings reflect a genuinely small underlying effect rather than insufficient sensitivity. The Bayesian framework provides direct quantification of the magnitude and uncertainty of treatment effects. Posterior point estimates for the primary Group × Time interactions were close to zero across all analyses with Crls. Nevertheless, modest sample sizes still limit our ability to characterise subgroup-level heterogeneity, which is particularly relevant given the growing view that group-level EEG differences in ADHD may have limited utility when used as an individual-level biomarker (Michelini, Norman, et al., 2022). From this perspective, the absence of differential change does not preclude clinically meaningful benefits in a subset of participants, but suggests that future studies may need to focus on individual-level prediction and treatment-by-biomarker interactions rather than average power shifts.

The TOVA task itself imposes constraints. The fixed TOVA structure confounds time-dependency with the non-counterbalanced H1/H2 switch in target frequency, and the task is not designed to elicit large numbers of errors, which left too few error trials for reliable spectral comparisons between correct and error responses (Cowley et al., 2022).

We also did not individualise the alpha band based on each participant’s individual *α* peak frequency (iAF), which may have reduced sensitivity to detect *α*-related effects. Recent evidence showed that, in a neurotypical group, NFB-induced increases in iAF causally enhanced attentional performance through accelerated *α* desynchronization dynamics (Jacques et al., 2026). Nevertheless, iAF-based banding was not adopted in the present study as it can introduce additional filtering-related confounds in trial-wise analyses (Cowley et al., 2016; Lansbergen et al., 2011).

In summary, extending our approach in future work should ideally extend TOVA itself, with condition counterbalancing and targeted diagnostic component-specific contrasts, which would necessitate re-recording the task’s normative databases (Greenberg et al., 2016).

Finally, follow-up duration between conclusion of treatment and outtake was six months; in future work, both immediate and one year or longer follow-ups could be added to assess how neurophysiological changes evolve over time.

### Summary

In summary, our results demonstrate that using standardized NFB training protocols as nonpharmacological treatment did not produce measurable improvements in sustained attention in adults with ADHD. Across behavioural performance (TOVA), attention-related ERPs (N2/P3), and spectral dynamics (*θ*/*α* power), we found no evidence for treatment-specific effects when comparing the treatment group with waiting-list controls, and no evidence for any difference in how ADHD groups changed following treatment with respect to neurotypical controls. The consistency of null findings across these complementary measures strengthens confidence that NFB did not meaningfully engage or modify the neurocognitive processes underlying attention deficits in adult ADHD.

## Materials and Methods

### Participants

From an initial sample of 80 volunteers, we screened (see below) and recruited 54 adults (25 males, *M_age_* = 36.26, *SD* = 10.22) diagnosed with ADHD. We then recruited an age-matched neurotypical control sample of 18 adults (6 males, *M_age_* = 32.78, *SD* = 10.82) with no diagnosed neurocognitive deficits or ongoing medication for ADHD/ADD. For more details of the RCT design, and the results derived from it, see previously reports by Cowley et al. (2016, 2022).

For screening, inclusion criteria for the ADHD group were (1) a diagnosis of ADHD/ADD, (2) no neurologic diagnoses, (3) age 18–60 years, (4) scores on ASRS (Kessler et al., 2005); and the Brown ADHD Scale (BADDS; Brown, 1996), indicating the presence of ADHD (descriptive statistics reported in Cowley et al. (2022), and (5) an IQ score of at least 80, measured by a qualified psychologist using Wechsler Adult Intelligence Scale IV. No strict cut-off values were used for ASRS and BADDS to indicate the presence of ADHD. Regarding exclusion criteria, the consulting psychiatrist decided exclusions based on structured clinical interviews with participants, following the guidelines of the Diagnostic Interview for ADHD in Adults (DIVA 2.0; Ramos-Quiroga et al., 2019). Several scales were used to evaluate comorbidities during the clinical interview. These included extreme outlier scores on the scales for Generalized Anxiety Disorder (Spitzer et al., 2006), the Beck Depression Inventory (Beck et al., 1996), the Alcohol Use Disorders Identification Test (Saunders et al., 1993), the Mood Disorder Questionnaire (Hirschfeld et al., 2000), the test of prodromal symptoms of psychosis (Heinimaa et al., 2003), and the Dissociative Experience Scale (Liebowitz, 1992) for dissociative symptoms. The psychiatrist made a final assessment of the balance of symptoms contributing to the patients’ presentation.

Outtake measurement was conducted in a between-subject manner using the waiting-list control design (WLC). The WLC design placed a randomly assigned ADHD participant on hiatus as ADHD-W group, while active treatment was applied to the randomly assigned treatment group (ADHD-T). Thus, participants in the intake ADHD group were divided equally between ADHD-T (n=27) and ADHD-W group (n=27); however, two switched from the ADHD-T group (final n=25) to the ADHD-W group (final n=29) for personal reasons. From these assignments, eight dropped out of the ADHD-W group (including one participant whose pretest measurement data was then deleted by request), and two from the T group. Thus, 23 in the ADHD-T group and 21 in the ADHD-W group (23 males, *M_age_*=38.59, *SD*=10.27) were available for analysis. The groups did not differ in terms of age, gender, or handedness. Because of reasons of data quality (detailed in Section: Epoch Extraction and Trial Selection) a number of participants were dropped from EEG analysis for different TOVA conditions. The final analysed number of participants were: at intake, ADHD inhibition H1 41, H2 36; response H1 41, H2 39; neurotypical control inhibition H1 15, H2 12; response H1 14, H2 14. At outtake, ADHD-T inhibition H1 21, H2 21; response H1 21, H2 21; ADHD-W inhibition H1 20, H2 19; response H1 20, H2 20.

### Procedure

The behavioural and EEG data were measured in a 2.5-to 3-hour multitask session, gathered in an electrically shielded and sound attenuated room. The full session included preparation (30-40 min), pretest baseline measurement (5 min), TOVA (22 min), vigilance measurement in the resting state (20 min) (Olbrich et al., 2012), a second novel CPT (∼20 min) (Cowley, 2014), and baseline measurement after the test (2 min). The TOVA test was recorded toward the beginning of the measurement session as suggested by (Greenberg et al., 2016, p. 24). To control pharmacological confounds, participants with ADHD were required to withhold taking their medication for at least 48 hours (washout period) and all participants were asked to refrain from taking any stimulants (e.g., coffee, cigarettes, energy drinks) containing caffeine and nicotine. Karolinska sleepiness scale (KSS, Åkerstedt & Gillberg, 1990) was employed before recordings to control participants’ sleepiness levels.

### TOVA

We administered TOVA visual version 8, consisting of the test software, USB relay hardware, a low-latency micro switch hardware response button and Synchronization Interface hardware for test-to-EEG amplifier synchrony (all products of the TOVA Company). TOVA is a computerized performance test (CPT) with a monotonous hybrid Go/Nogo target-classification task design (Leark et al., 2004). For detailed task schematic and trial structures, see Figure 4. Participants were instructed to respond to the targets as accurately and quickly as possible and not to respond when they saw nontargets. A practice test of approximately 20 trials was administered before the actual test. Trials can be correct responses, commission errors (incorrect responses), omission errors (incorrect inhibition), and correct inhibition. TOVA has two consecutive equal-length conditions: the infrequent and frequent target modes. During the H1, the target stimulus appears infrequently (72 targets, and 252 nontargets, i.e. 22.5% target probability), which constitutes a test of vigilance and a challenge to inattentiveness. During the H2, these frequencies are reversed (252 targets and 72 nontargets, i.e. 77.5% target probability), which constitutes a test of inhibition and a challenge to hyperactivity. Participants are not informed about this transition. Each stimulus is displayed for 100 ms and is separated from the next stimulus by a 2000 ms inter-stimulus interval. Following the physiological constraints of human reaction times, RTs *<* 150 ms were classified as anticipatory responses and excluded (Whelan, 2008). The duration of TOVA is 21.6 minutes, with each condition lasting 10.8 min.

**Figure 4:**
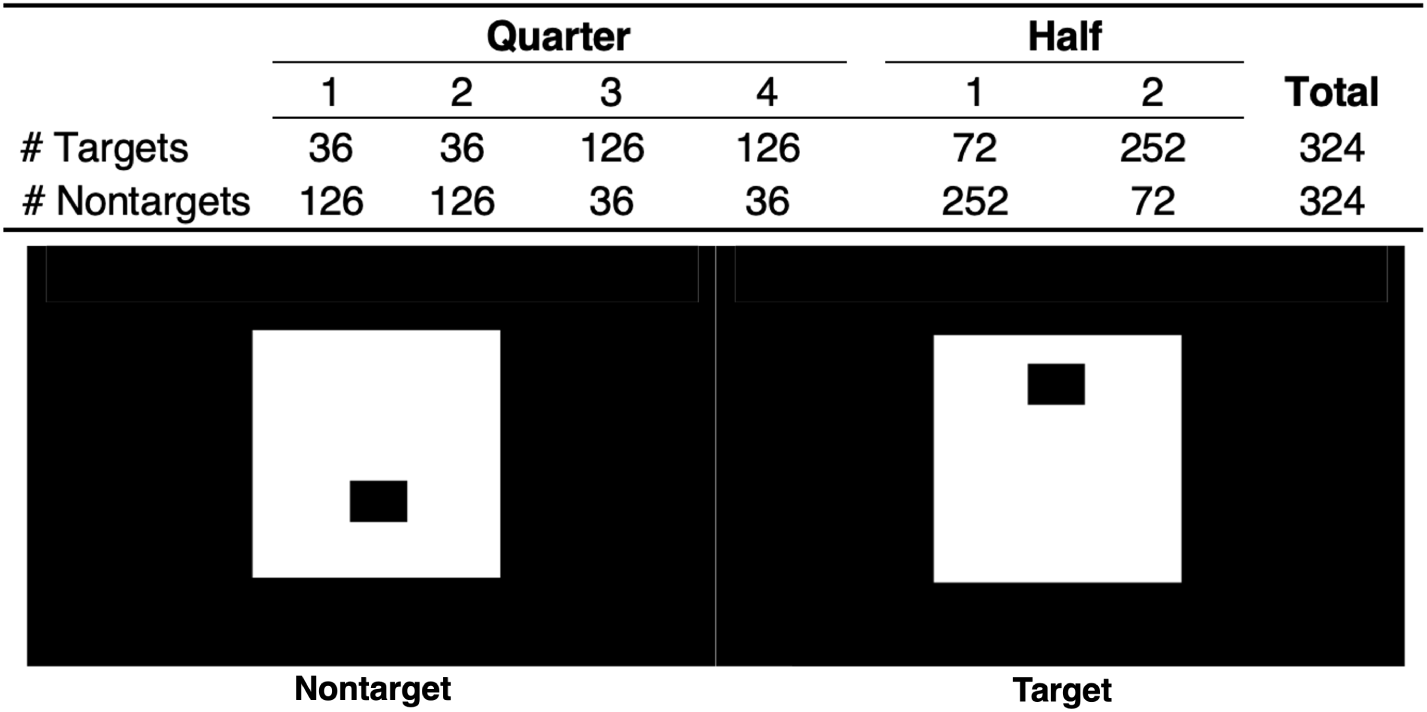
TOVA experiment protocol design. Top panel: Number of targets and nontargets in each quarter and half, illustrating target-to-non-target frequency. Bottom panel: Target and non-target stimulus characteristics: a white rectangle on a black screen, with a smaller black rectangle in one of two offset positions denoting trial type.

### TOVA Variables

Analysis was conducted on standardized TOVA scores, derived by comparing individual performance against a normed sample population (N=1596), this implies that behavioural variables do not represent, for example, actual error but rather the difference from a ‘normal’ value for error (Greenberg et al., 2016, p. 24). The comparison group for each participant is determined by their age group and gender. TOVA software classifies the performance of the test subjects as ‘not within normal limits’, ‘borderline’, or ‘normal’. Five standard scores are computed: (1) mean response time (MRT): mean of correct RTs in milliseconds (ms); (2) RTV: SD of mean correct RT; (3) commission errors: incorrect responses to a nontarget stimulus; (4) omission errors: incorrect responses to a target stimulus; and (5) *d^′^*: the ratio of hits (correct responses) to false alarms (commission errors). Since the five variables are standardized by normative comparison, where 100 is the reference value, then one should interpret them such that values lower than 100 indicate more error, more RTV, etc.

TOVA administration is recommended between 6:00 and 13:00 hours. Measurement times at intake were close to or within the normed hours, as detailed in Cowley et al. (2022). At outtake, ADHD-T group *M* = 14:45, *SD* = 2 h 3 min; ADHD-W group *M* = 14:21, *SD* = 2 h 17 min. The TOVA starting time did not differ statistically between the groups (*t*_(40)_ = 0.623, *p* = 0.537).

### EEG Measurement and Analysis

EEG was measured using Biosemi Active Two equipment with 128 active electrodes mounted on a cloth headcap at equiradial positions. Active electrode CMS (Common Mode Sense) and passive electrode DRL (Driven Right Leg) were used to create a feedback loop for amplifier reference. Electrooculography (EOG) was recorded using a bipolar montage: horizontal EOG electrodes were attached to the outer canthi of both eyes; vertical EOG electrodes were attached above and below the left eye. Electrode offsets (running average of voltage at each electrode) were kept below ±25 *µ*V. During TOVA, participants were advised to relax, sit as still as possible, avoid excessive movements, and fixate on the middle of the screen, which was shown as a small white dot between target stimulus presentations.

### Preprocessing

The data underwent preprocessing with the Computational Testing Automated Preprocessing Toolbox (CTAP; Cowley & Korpela, 2018; Cowley et al., 2017) based on EEGLAB for MATLAB (Delorme & Makeig, 2004). Offline re-referencing was performed to the average of the two mastoids. A low-pass filter at 45 Hz and a high-pass filter at 2 Hz were applied. CTAP was used to detect eye blinks using the probabilistic method in the time domain described in Cowley et al. (2017), whose validity was examined for each participant by visual inspection.

Continuous EEG and EOG data for each participant were decomposed using the FastICA algorithm to identify and remove ocular and muscular artifacts (Hyvärinen, 1999). Independent components (ICs) statistically similar to CTAP-detected blinks were removed (Cowley et al., 2017). In addition, artefactual channels, epochs, and ICs were identified using fully automated statistical thresholding for the rejection of EEG artifacts (FASTER; Nolan et al., 2010). The automatically rejected channels were interpolated from the adjacent electrodes. Fluctuations exceeding ±80 *µ*V in amplitude across 50% of channels were discarded as bad segments. The remaining ICs of ocular or muscular artifact were manually rejected by joint visual inspection, using the ICLabel function in EEGLAB (Pion-Tonachini et al., 2019). On average, 3% of channels and one IC per participant were rejected at intake, and 5% of channels and 22% of ICs per participant were rejected at outtake.

All subsequent analyses were focused on 2 regions-of-interest (ROIs), frontal and parietal, with 8 channels each denoted using Biosemi 128 channel labels as: frontal C4, C13, C14, C19, C21, C26, C27, D4, and parietal A4, A5, A7, A17, A21, A30, A32, B4. Mapping from Biosemi 128 into International 10–5 electrode placement system (Jurcak et al., 2007), by selecting closest matches in terms of angle theta/azimuth phi offset, the labels are: frontal at F4, F4h, Fz, F3h, F3, AFF4h, AFz, and AFF3h and parietal at P3, P1, CPPz, P2, P4, PO3, POz, and PO4. The electrode locations were chosen *a priori* based on previous research (Hanslmayr et al., 2005, 2007; Macdonald et al., 2011; Mathewson et al., 2009).

### Epoch Extraction and Trial Selection

For event-related analyses, the EEG data were extracted into four types of epochs: commission errors, correct responses, omission errors and correct inhibition. The epochs lasted 2000 ms, with 1000 ms before and after stimulus onset. Despite prior ICA-based blink removal, epochs that still contained a residual blink (as detected by CTAP) starting 400 ms before stimulus onset to 600 ms after stimulus onset were rejected. After this rejection, too few error trials remained to conduct reliable statistical analyses. At outtake, ADHD-W had on average 11 and ADHD-T had 10 commission error trials; ADHD-W had 4 and ADHD-T 12 omission error trials. At intake, controls had on average 7 and the combined ADHD group had 9 commission error trials; controls had 2 and ADHD had 6 omission error trials (Cowley et al., 2022). Thus, we analysed only the correct inhibition and response trials, in H1 and H2, to give four datasets per group for most analyses.

To counterbalance fluctuations in voltage offset across epochs and participants, a baseline correction was applied to each epoch, by subtracting the baseline mean amplitude from the entire epoch. The baseline period was from 1000 to 0 ms before stimulus onset.

In the intake EEG analyses (Cowley et al., 2022), a participant was only included if they had at least 36 epochs for infrequent conditions (responses H1 and inhibits H2) or 72 epochs for frequent conditions (inhibits H1 and responses H2) for each of these four conditions. This ensured each participant contributed 108 clean epochs per test half (i.e., H1 had 36 response epochs and 72 inhibit epochs, totalling 108 epochs, and vice versa for H2). However, we slightly adjusted the trial-inclusion approach by applying a limited amplitude-based bootstrapping procedure, in order to retain more participants in the pre–post EEG analyses and thereby increase statistical power. For each group × condition × half, we computed the group-level distribution of mean absolute amplitude across all correct trials within the conditions. Participants with fewer than 80% of the target number of epochs were excluded. For participants whose epoch counts were close to the threshold (at least 80% but still below the 36/72 threshold), we imputed the missing epochs by resampling with replacement from their existing epochs whose mean absolute amplitude was closest to the group-level mean. This procedure was required simply because participant-wise data must all be of same dimension, and allowed participants with near-complete data to contribute to the analyses. Additionally, to address potential biases influenced by the sampling process, the epochs were randomly sampled without replacement from the total available per condition on a participant-wise basis. To test that this sampling did not bias results, we conducted 11 similar draws and estimated the variance across draws of the spectral power, following the same approach in (Cowley et al., 2022). Mean variance was 0.015 for intake and 0.026 for outtake (averaged across baseline and post-stimulus periods).

### ERP Calculation

Grand-average ERP waveforms were computed for each group (ADHD-T, ADHD-W) at each time (intake, outtake) within frontal and parietal ROIs. Data were epoched to 1-s windows time-locked to target onset. Additionally, ERP images displaying trials individually (amplitude visualized with colour, sorted by RT) at outtake were generated for supplementary materials, following the approach described in Cowley et al. (2022). These images were generated by ordering the trials according to their RT value and smoothing across the trials with a moving Gaussian of width proportionate to the number of trials (Gaussian *SD* = trial number/30 ⇒ window width = trial number/5).

### Spectral Power Calculation

To analyse how the EEG spectrum changes from pre-stimulus to post-stimulus, we calculated oscillatory power separately for these periods. This was done using Welch’s PSD estimation through the EEGLAB spectopo() function. The Fourier transform window length was set to 512 (matching the sample rate) with an overlap of 384 frames. Power was calculated for each 1-Hz frequency bin from 4 to 16 Hz. We also examined the pre-stimulus activity. For this, power was calculated for the 500 ms pre-stimulus period, focusing on parietal ROIs. Correct inhibition and correct response trials were included because this analysis specifically focused on pre-stimulus activity. Within these trials, the *α* power was calculated, focusing on 9–12 Hz.

### Statistical Analysis

#### Behavioural Performance

Cowley et al. (2016) initially reported null treatment effects using independent-samples *t*-tests. We re-analysed the behavioural data using Bayesian linear mixed model (LMM; brms package in R; Bürkner, 2017). Compared with the fixed-effect models used for the intake analyses in Cowley et al. (2022), the Bayesian approach is better suited to the present repeated-measures design (Hesser, 2015; Yu et al., 2022). We then fitted a multivariate model analysing all five variables (MRT, RTV, commission errors, omission errors, and *d^′^*). The model specified fixed effects for Group (ADHD-T vs. ADHD-W), Time (intake and outtake), TOVA condition (H1: infrequent targets; H2: frequent targets), and their three-way interaction (Group × Time × TOVA condition). To account for potential individual differences at baseline, TOVA’s *Attention Performance Index* (API) was included as a covariate (Leark et al., 2004). Random effects included random intercepts and random slopes for Time by participant. Models used weakly informative priors and a Student-*t* likelihood to improve robustness to skewed and heavy-tailed distributions. Posterior distributions were estimated using four Markov chain Monte Carlo (MCMC) chains with 12,000 iterations each (3,000 warm-up). We report posterior means (*β*) and 95% credible intervals for all fixed effects. We assessed convergence and sampling diagnostics for Bayesian analyses by inspecting *R*^^^, divergent transitions, and also by conducting posterior predictive checks. All Bayesian models converged well (*R*^^^≤1.007). Detailed diagnostics are provided in Supplementary Table S5.

Potential extreme outliers were assessed by examining studentized residuals exceeding ±3. We note, however, that excluding outlier scores in a clinical sample is not straightforward, as high behavioural variability itself is a clinical characteristic in adults with ADHD (Castellanos et al., 2005). In total, nine outlier scores were excluded from the analyses of standardised scores, and removing these values did not change the interpretation of the results. The case list of excluded outliers are reported in Supplementary Table S1. In order to validate that the neural data analysis would not be confounded by relative variation in motor processing activations, due to any observed between-group difference in the number of button presses, we conducted the following analysis. We tested whether there was a difference in the total number of button presses (= correct target presses + commission errors) between groups or test quarters (Q1–Q4) by fitting a robust LMM (Koller, 2016) with total presses as the dependent variable (DV), and group (ADHD-T vs. ADHD-W), time (intake and outtake), test quarters, and their interactions as predictors. A numerical participant ID was used as a random factor, allowing variability in the intercepts but not the slopes. Robust LMM was employed because total button presses were bimodally distributed across test quarters. Across both intake and outtake, there were substantially fewer presses during Q1–Q2 (infrequent target, *M* = 36.3) than during Q3–Q4 (frequent target, *M* = 127.9). Neural results were not confounded by between-group differences in motor processing (see Supplementary Materials).

Additionally, we fitted a Bayesian LMM comparing each ADHD group against the neurotypical control (NTC) group at *intake* and *outtake* to characterize behavioural differences. This model followed the same specifications as described above. Full results are reported in Supplementary Figure S1.

### EEG Data

Statistical analysis of EEG was performed in MATLAB (version 9.17) using the EEGLAB toolbox (Delorme & Makeig, 2004), and in the R platform for statistical computing (R Core Team, 2020).

### ERP analysis

Group differences in correct response ERPs were tested using Bayesian LMMs of the mean amplitude and peak latency within two predefined windows in target-locked trials: 150-250 ms for the N2 component and 330-430 ms for the P3 component in target-locked trials. These ERP windows were defined based on the grand-average ERP waveforms across all ADHD participants and healthy controls at intake, as reported in (Cowley et al., 2022). These windows were centred on the N2 and P3 waves observed in the data. Using baseline ERP windows for pre-and-post analysis follows standard practices, avoiding circularity while ensuring consistency with our previously published baseline findings (Luck, 2014). Visual inspection of outtake waveforms confirmed that N2 and P3 components remained within these predefined windows (see Supplementary Figure S1, gray shaded areas). Each Bayesian LMM included the three-way interaction of group × measurement time × ROIs, with TOVA condition was included as a covariate. Random effects included random intercepts and random slopes for measurement time by participant ID. Models used weakly informative priors based on observed distributions and Student’s *t* likelihood for robustness. We focused on the Group × Time interaction to evaluate differential treatment effects between the groups. Additionally, we examined group differences at outtake as extra exploratory analyses by using ANOVA with false discovery rate (FDR) correction for multiple comparisons (Supplementary Figure S3). We also fitted two Bayesian LMMs comparing each ADHD group against the NTC group at *intake* and *outtake* to characterize ERP differences in mean amplitude and peak latency at N2 and P3 windows. These models followed the same specifications as described above. Full results are reported in Supplementary Figure S2.

### Spectral analysis

We analysed the differences between baseline and post-stimulus periods in their log-PSD at 4- and 8-Hz frequency bins (these frequencies were chosen based on their role in attention within TOVA, and confirmed by visual inspection at intake). We fitted two separate Bayesian LMMs for the 4 Hz and 8 Hz frequencies, with log-PSD as the DV. Each model included the four-way interaction of period (baseline, post-stimulus) × measurement time × group × ROI, with TOVA condition was included as a covariate. Random effects included random intercepts and random slopes for measurement time by participant ID. Models used weakly informative priors and Gaussian likelihood. To examine the potential differences of parietal pre-stimulus *α* power (8–12 Hz), a separate Bayesian LMM was fitted to examine the group × measurement time inter-action in parietal pre-stimulus *α* power on correct trials, with TOVA condition included as a covariate. All Bayesian LMMs were estimated using 4 MCMC chains with 12,000 iterations (3,000 warmup).

## Data Availability

The data and analysis code that support the findings of this study are available from the corresponding author upon reasonable request. Raw EEG data are not publicly available due to privacy restrictions related to clinical trial participants under the European General Data Protection Regulation (GDPR).

## Acknowledgements

The authors wish to thank Guang Rong who provided suggestions on statistical analyses, and Kristiina Juurmaa for her previous work in EEG data gathering and analysis.

## Additional information

### Author contributions

Conceptualization: BC. Data curation: BC, JW, AR. Formal analysis: JW, AR, BC. Investigation: JW, BC. Methodology: BC, JW, AR. Resources: BC. Software: JW, BC. Supervision: BC, AR. Visualization: JW. Writing – original draft: JW. Writing – review and editing: AR, BC.

## Footnotes

1 Inverse training was a novel addition of a self-regulatory component designed to address the heterogeneity and aetiological uncertainty of ADHD in the adult population (Veilahti et al., 2021).

2 Note: because these scores are standardised, they must be interpreted in terms of deviations from the reference level of 100: for example, 85 thus means ∼1SD below the norm.

3 Note: interpretation of TOVA behavioural data should bear in mind the interaction of variables, and consider the full profile, not one score alone. For example, excessive commissions or anticipatory responding can artificially shorten response time and affect variability (Greenberg et al., 2016).

