## Supplementary Material for "Neural and behavioural measures from attention testing show no support for efficacy of neurofeedback treatment for adult ADHD"

Neurofeedback treatment for adult ADHD produces no effect on TOVA variables – Supplementary Materials

### Behavioural Performance

| **Table S1:** **TOVA behavioural observations identified as potential extreme outliers** | | | | | | |
| --- | --- | --- | --- | --- | --- | --- |
| **Variable** | **Participant ID** | **Group** | **Time** | **TOVA condition** | **API** | **Score** |
| MRT | 1065 | ADHD-T | *Outtake* | H2 | -14.90 | 16.32 |
| RTV | 1065 | ADHD-T | *Intake* | H2 | -9.62 | -44.76 |
|  | 1036 | ADHD-T | *Outtake* | H1 | -17.45 | -155.15 |
|  | 1081 | ADHD-T | *Outtake* | H1 | -11.97 | -100.50 |
| Commission Errors | 1075 | ADHD-W | *Outtake* | H1 | -10.34 | -304.82 |
| d-prime (*d'*) | 1068 | ADHD-W | *Outtake* | H2 | 9.57 | 191.70 |
| Omission Errors | 1036 | ADHD-T | *Outtake* | H2 | -17.45 | -1249.41 |
|  | 1058 | ADHD-T | *Outtake* | H2 | -15.10 | -1174.44 |
|  | 1065 | ADHD-T | *Outtake* | H2 | -14.90 | -1437.75 |
| ***Note.*** Scores > 85 are within or above normal limits, scores 80-85 indicate borderline ADHD, score < 80 indicate performance that is not within normal limits. API: Attention Performance Index. | | | | | | |

| **Table S2: Descriptive statistics of key standardized variables across measurement times** | | | | | |
| --- | --- | --- | --- | --- | --- |
| **Variable** | **TOVA condition** | **ADHD-T** | | **ADHD-W** | |
|  |  | ***Intake*** | ***Outtake*** | ***Intake*** | ***Outtake*** |
| MRT | H1 | 106.82 ($\pm$13.30) | 104.63 ($\pm$18.14) | 109.96 ($\pm$15.62) | 107.37 ($\pm$14.55) |
|  | H2 | 106.94 ($\pm$16.57) | 103.21 ($\pm$23.33) | 111.28 ($\pm$16.91) | 107.23 ($\pm$20.64) |
| RTV | H1 | 89.56 ($\pm$21.53) | 69.92 ($\pm$70.24) | 86.49 ($\pm$37.71) | 81.99 ($\pm$34.91) |
|  | H2 | 79.51 ($\pm$38.82) | 78.47 ($\pm$51.47) | 91.29 ($\pm$23.05) | 81.11 ($\pm$41.25) |
| Commission Errors | H1 | 95.89 ($\pm$11.62) | 97.30 ($\pm$19.82) | 98.08 ($\pm$21.69) | 82.89 ($\pm$89.64) |
|  | H2 | 78.55 ($\pm$29.10) | 86.79 ($\pm$25.52) | 81.98 ($\pm$23.06) | 87.47 ($\pm$28.83) |
| Omission Errors | H1 | 100.12 ($\pm$4.60) | 85.64 ($\pm$36.75) | 95.56 ($\pm$15.04) | 94.26 ($\pm$15.22) |
|  | H2 | 61.90 ($\pm$78.54) | -93.27 ($\pm$475.39) | 70.12 ($\pm$83.43) | 25.02 ($\pm$183.43) |
| d-prime (*d'*) | H1 | 89.40 ($\pm$17.62) | 90.54 ($\pm$29.12) | 92.45 ($\pm$22.11) | 96.30 ($\pm$19.87) |
|  | H2 | 71.31 ($\pm$34.21) | 74.86 ($\pm$43.83) | 79.87 ($\pm$21.55) | 84.42 ($\pm$42.01) |
| ***Note.*** Scores > 85 are within or above normal limits, scores 80-85 indicate borderline ADHD, score < 80 indicate performance that is not within normal limits. Values in the table are presented as Mean ± Standard Deviation (SD). The large SD and negative mean observed in omission errors (ADHD-T) at *outtake* reflect the inclusion of valid but abnormal performances. | | | | | |

### Motor Processing Control Analysis

In the analysis on differences in total number of button presses, we fitted a robust LMM. The Group Time interaction was not significant (β = 0.29, *SE* = 0.95, *t* = 0.30), indicating no differential change in total button presses between the groups from *intake* to *outtake*. Similarly, all Group × Time × Test Quarters interactions were all non-significant (all |*t*| < 0.5). Estimated marginal means (averaged across quarters) showed minimal group differences at both *Intake* (ADHD-T: 83.0 presses, ADHD-W: 83.1; difference = -0.11, *p* = 0.818) and *Outtake* (ADHD-T: 82.3, ADHD-W: 82.6; difference = -0.27, *p* = 0.607). Thus, the neural results were not confounded by between-group differences in motor processing activations.

### ADHD groups relative to neurotypical control group

#### Behavioral Performance

To contextualize TOVA behavioral performance relative to neurotypical control (NTC) group, we fitted a Bayesian LMMs comparing ADHD-T and ADHD-W *intake* and *outtake* against the NT group. DVs were standardized prior to modelling, so estimates represent standardized posterior mean differences. The results are shown in Figure S1 below.

For ADHD-T, the only credible difference from NT was observed for *d’* during H1 condition at *intake*, with ADHD-T showing lower standardized scores than NT, estimate = -0.381, 95% Crl [-0.670, -0.101]. No credible ADHD-T differences from NT were observed at *outtake*, and all other ADHD-T contrasts included zero. For ADHD-W, credible differences from NT were also observed for H1 *d’* at *intake*, estimate = -0.283, 95% Crl [-0.556, -0.004], and H2 commission error scores at *intake*, estimate = -0.417, 95% Crl [-0.797, -0.048]. At *outtake*, ADHD-W showed lower H1 MRT, estimate = -0.469, 95% Crl [-0.870, -0.076], and lower H1 RTV, estimate = -0.396, 95% Crl [-0.634, -0.162], relative to NT. No credible differences were observed for omission errors.

Overall, several *intake* differences from NT were attenuated at *outtake*, most clearly for H1 *d’* in both ADHD groups and H2 commission errors in ADHD-W. However, this pattern was not consistent across all DVs, as ADHD-W showed larger negative differences from NT at *outtake* for MRT and RTV during H1.

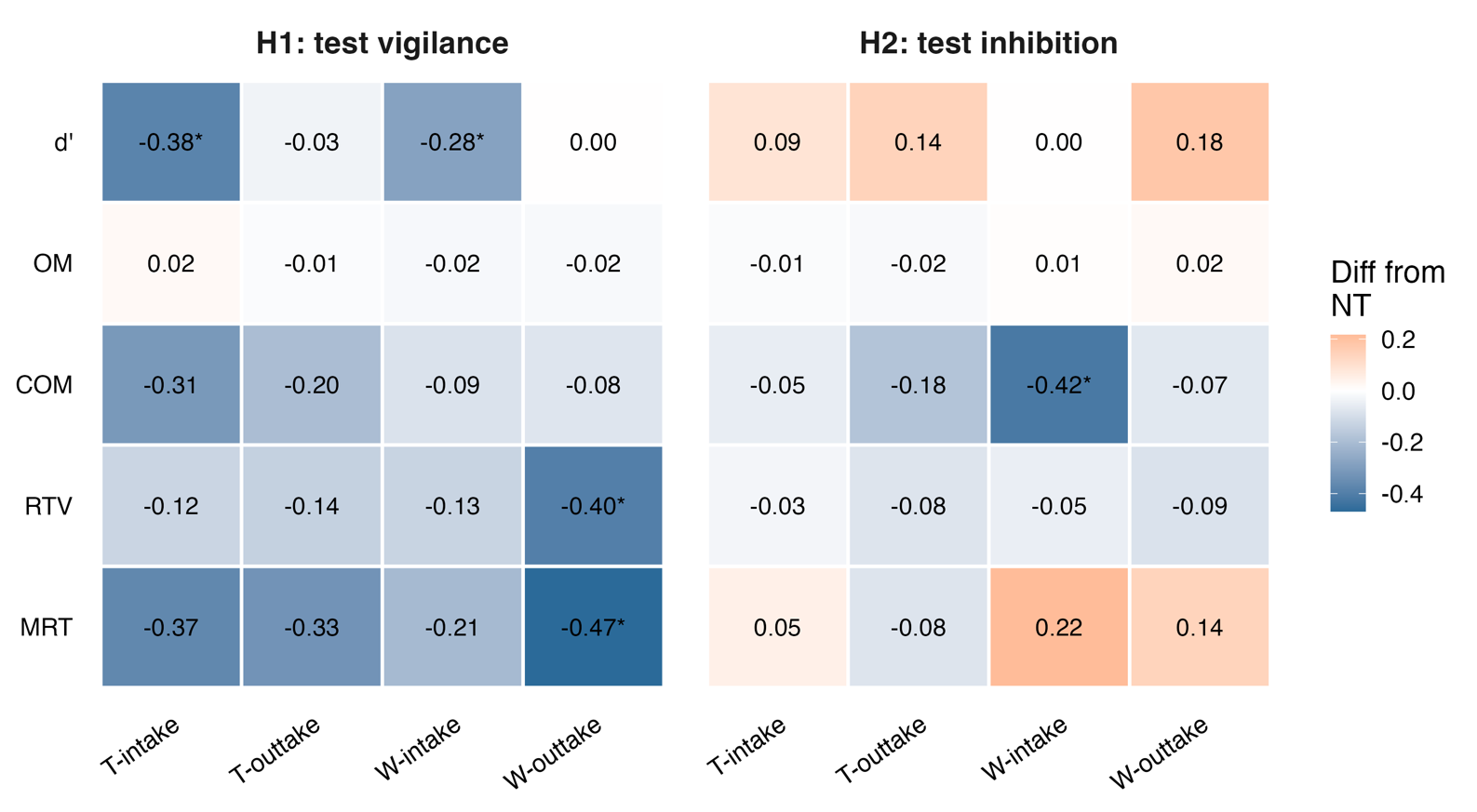

**Figure S1. Posterior contrasts of behavioral performance relative to neurotypical controls.** Heatmap values represent posterior mean standardized differences between each ADHD condition and the NT group. Negative values indicate lower standardized scores relative to NT; asterisks indicate 95% Crl excluding zero.

#### ERP Analysis

To further understand ERP patterns relative to the NT group, we fitted two Bayesian LMMs for ERP mean amplitude and peak latency, comparing the two ADHD groups at *intake* and *outtake* against the NT group. The results are shown in Figure S2a and Figure S2b below.

For mean amplitude, both ADHD groups showed consistently reduced frontal P3 amplitudes relative to NT, with credible differences emerging particularly at *outtake*. At parietal sites, both ADHD groups exhibited reduced N2 negativity relative to NT, most prominently during H1.

For peak latency, differences relative to NT were generally small and mostly non-credible. The only contrast reaching the 95% credible interval threshold was a delayed parietal P3 latency during H1 in ADHD-W at *intake*. No credible N2 latency differences were observed, and the remaining latency contrasts did not show a consistent pattern across ROIs, task halves, or timepoints.

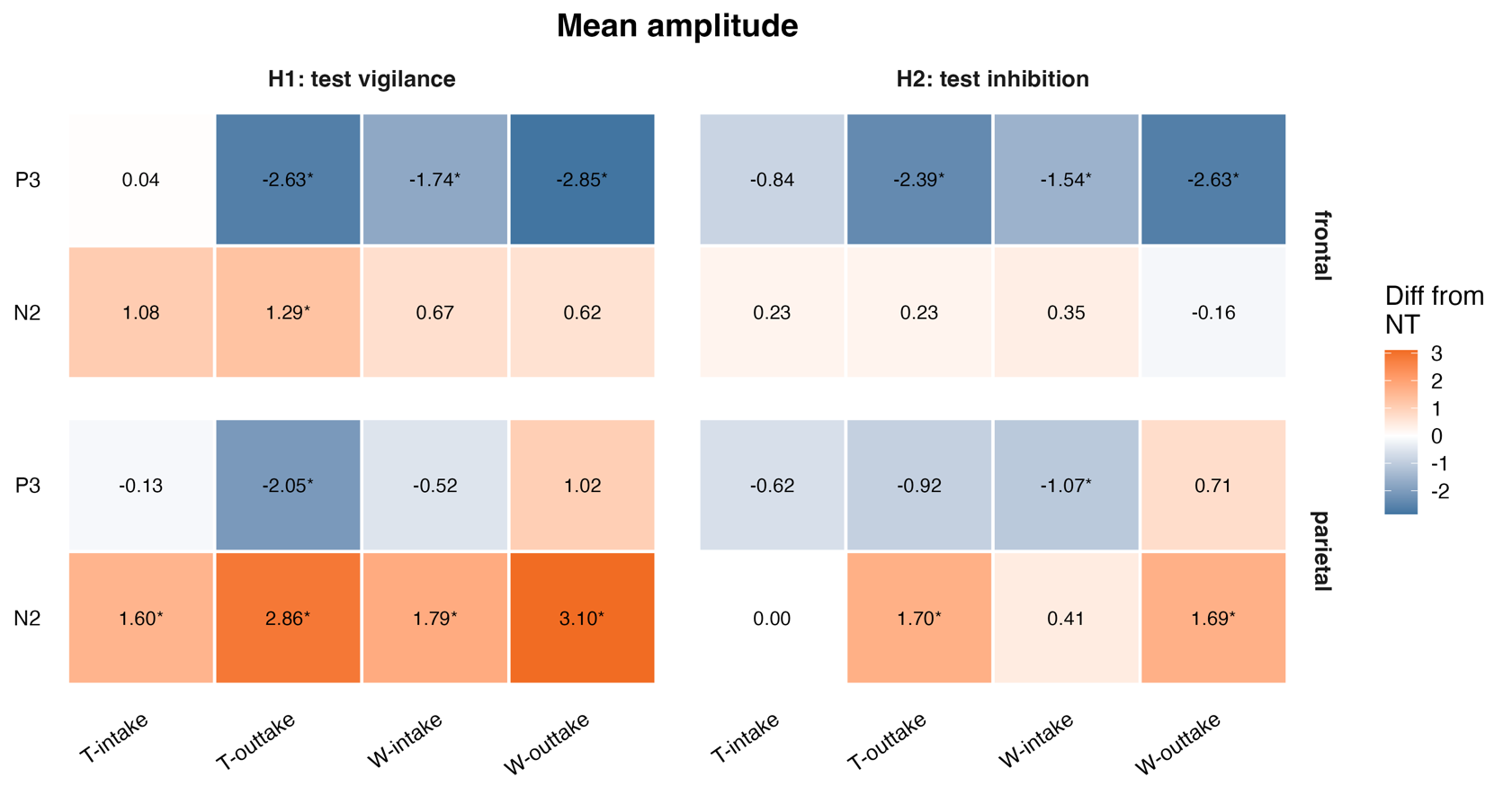

**Figure S2. Posterior contrasts of ERP amplitude (above) and latency (below) relative to neurotypical controls.** Heatmap values represent posterior mean differences (amplitude: uV; latency: ms) between each ADHD condition and the NT group. Asterisks indicate 95% Crl excluding zero.
For N2 amplitudes, positive differences indicate reduced negativity. Positive/negative values indicate delayed/earlier latency relative to NT.

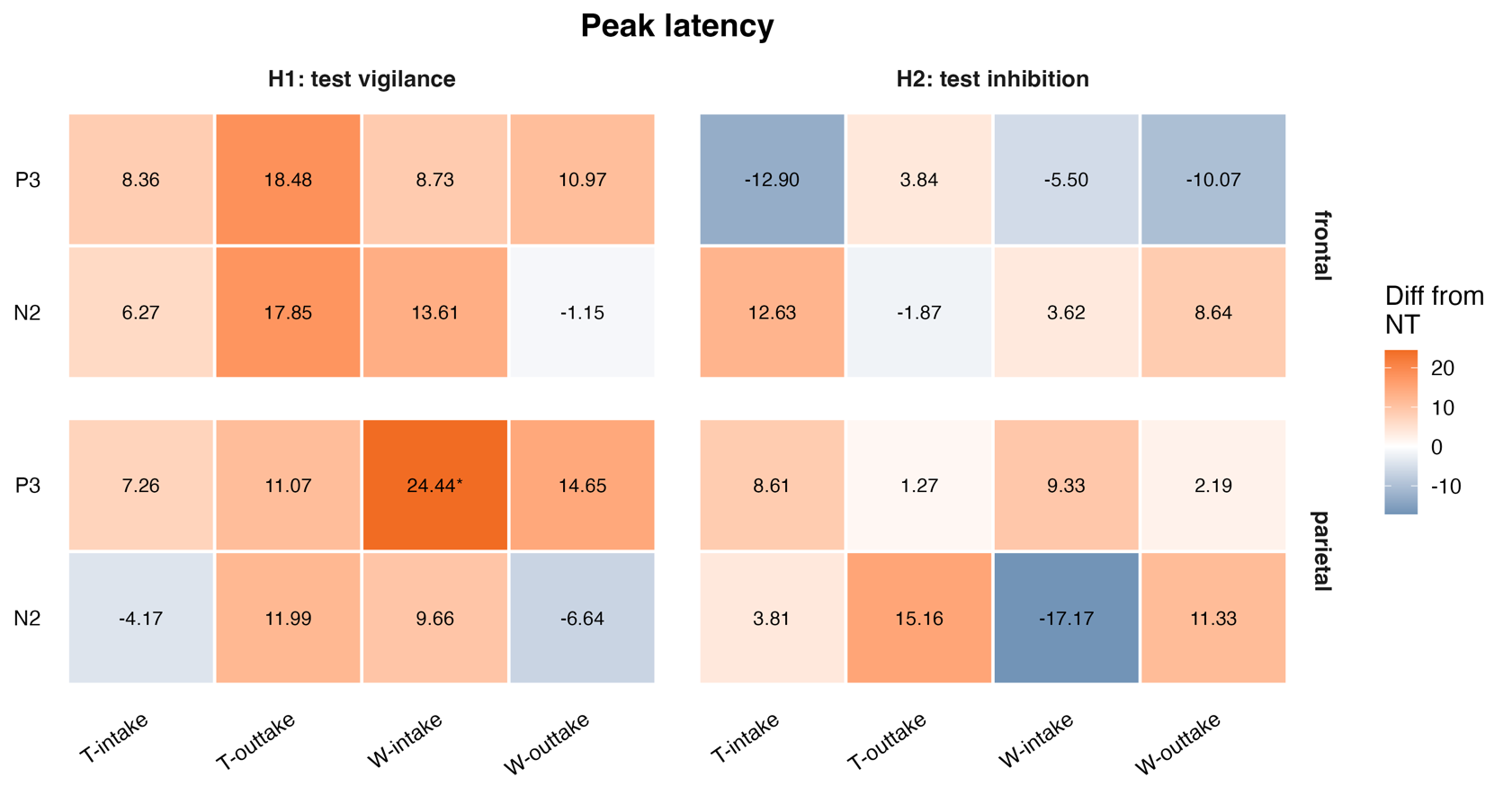

### Target- vs. Response-locked ERPs

The figures S3 below present RT-sorted ERP image plots from the *outtake*, illustrating group-wise differences between ADHD-T and ADHD-W in target-locked and response-locked amplitudes at frontal and parietal ROIs. Group differences in grand-average ERP waveforms were tested using ANOVA with FDR correction for multiple comparisons. The ADHD-T group exhibited significantly stronger frontal P3 amplitudes in the target-locked condition during both H1 (*p_adj_* <0.05) and H2 (*p_adj_* <0.0001), as well as significantly stronger negative potentials in the post-response window. Parietal N2 latency was significantly shorter in the ADHD-T group during H2 (*p_adj_* <0.001).

**Figure S3. RT-sorted ERP visualization in TOVA H1 and H2 in *outtake*.** Each row is a single trial with color-coded amplitude from -8 to +8 mV (cool and warm colors, respectively). On the left are target-locked trials from -200 to +800 ms of stimulus onset, and time of response is shown by the black sigmoidal curve. On the right are response-locked trials from -600 to +400 ms of RT, and stimulus onset is shown by the black sigmoidal curve. Cumulative ERP waves are shown below each ERP image, split by the median RT. First and second rows (both H1 and H2 panels). These ERP images show stimulus-locked early waves (i.e., amplitudes at a fixed lag from 0). Third row (both panels): ERP waves (solid lines) for each ROI. Vertical gray areas are test windows (aligned to N2, P3 in target-locked trials). Group differences are annotated as * *p_adj_* < 0.05, **** *p_adj_* < 0.0001.

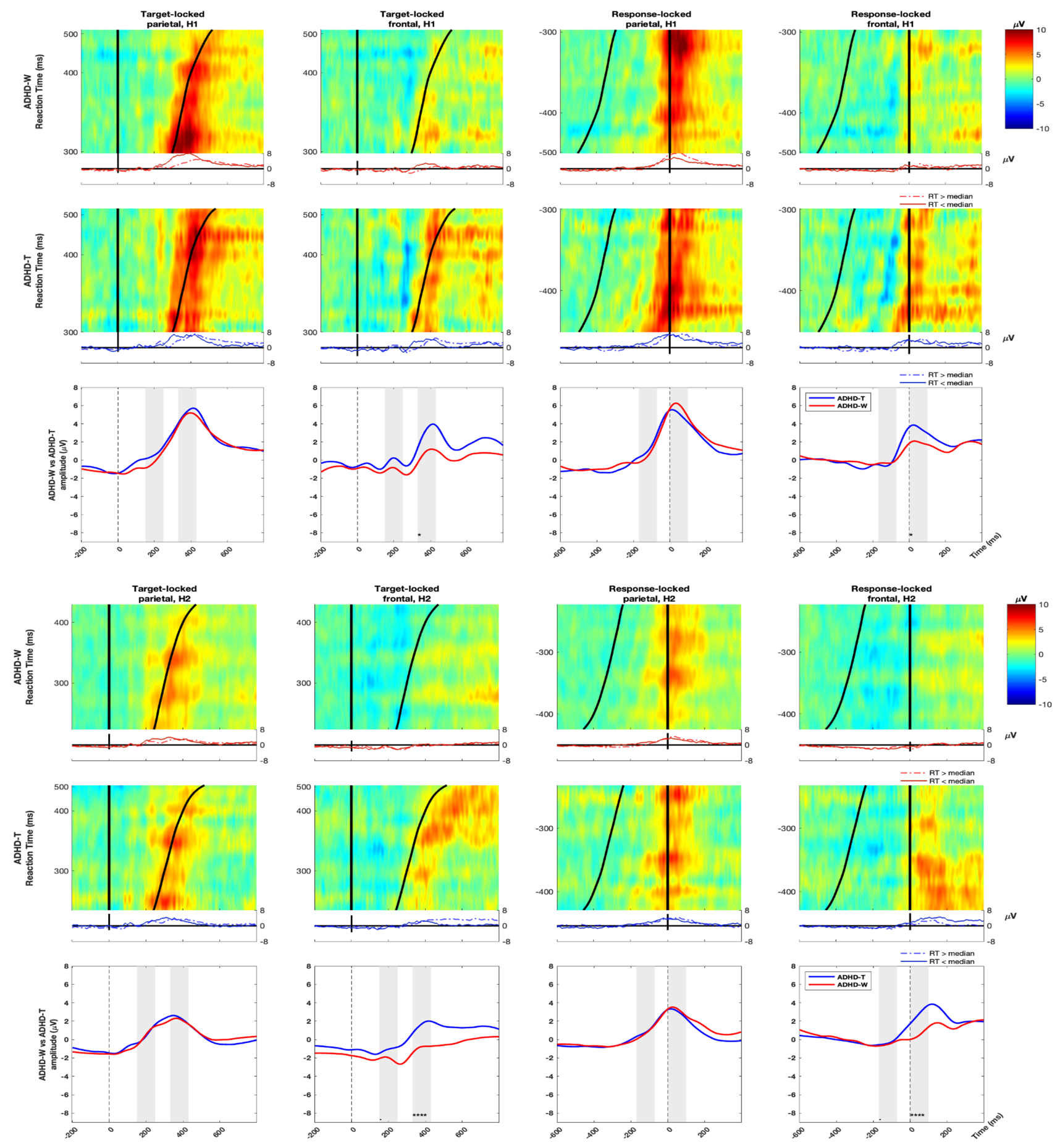

| **Table S3:**  **Bayesian linear mixed model results for ERP analysis** | | | | |
| --- | --- | --- | --- | --- |
| **Component** | **Measure** | **Parameter** | **Estimate** | **95% Crl** |
| **N2** | **Mean amplitude** | Intercept | -0.15 | [-0.78, 0.46] |
|  |  | H2 | -0.01 | [-0.17, 0.16] |
|  |  | Group (ADHD-W) | 0.24 | [-0.55, 1.02] |
|  |  | Time (*outtake*) | 0.01 | [-0.33, 0.34] |
|  |  | ROI (parietal) | -0.7 | [-1.00, -0.40] |
|  |  | Group × Time | -0.01 | [-0.42, 0.41] |
|  |  | Group × ROI | 0.62 | [0.23, 1.00] |
|  |  | Time × ROI | -0.02 | [-0.49, 0.44] |
|  |  | Group × Time × ROI | 0.02 | [-0.57, 0.61] |
| **N2** | **Peak latency** | Intercept | 212.67 | [201.70, 223.59] |
|  |  | H2 | -2.47 | [-8.31, 3.35] |
|  |  | Group (ADHD-W) | -12.08 | [-25.61, 1.76] |
|  |  | Time (*outtake*) | 0.87 | [-10.23, 11.96] |
|  |  | ROI (parietal) | -13.36 | [-22.77, -4.10] |
|  |  | Group × Time | -1.41 | [-15.78, 12.87] |
|  |  | Group × ROI | 18.54 | [6.53, 30.79] |
|  |  | Time × ROI | -1.58 | [-16.50, 13.27] |
|  |  | Group × Time × ROI | 2.38 | [-16.70, 21.41] |
| **P3** | **Mean amplitude** | Intercept | 2.37 | [1.74, 3.00] |
|  |  | H2 | -0.97 | [-1.16, -0.78] |
|  |  | Group (ADHD-W) | -0.43 | [-1.22, 0.38] |
|  |  | Time (*outtake*) | 0.01 | [-0.35, 0.36] |
|  |  | ROI (parietal) | -0.18 | [-0.47, 0.12] |
|  |  | Group × Time | -0.01 | [-0.47, 0.45] |
|  |  | Group × ROI | 0.1 | [-0.29, 0.49] |
|  |  | Time × ROI | -0.01 | [-0.49, 0.48] |
|  |  | Group × Time × ROI | 0.01 | [-0.63, 0.65] |
| **P3** | **Peak latency** | Intercept | 383.78 | [373.41, 394.08] |
|  |  | H2 | -5.74 | [-10.38, -1.14] |
|  |  | Group (ADHD-W) | -3.41 | [-16.56, 9.76] |
|  |  | Time (*outtake*) | -0.07 | [-9.02, 9.03] |
|  |  | ROI (parietal) | 5.03 | [-2.56, 12.56] |
|  |  | Group × Time | 0.09 | [-11.80, 11.89] |
|  |  | Group × ROI | -6.13 | [-16.09, 3.78] |
|  |  | Time × ROI | 0.27 | [-12.08, 12.58] |
|  |  | Group × Time × ROI | -0.4 | [-16.48, 15.73] |
| ***Note*:** Estimates are posterior means. Reference levels: Time = *intake*, Group = ADHD-T, ROI = frontal, Half = H1. | | | | |

### Spectral analysis

| **Table S4: Bayesian linear mixed model results for spectral power analysis** | | | | |
| --- | --- | --- | --- | --- |
|  | **Theta (4 Hz)** | | **Alpha (8 Hz)** | |
| Parameter | **Estimate** | **95 % Crl** | **Estimate** | **95 % Crl** |
| Intercept | 6.97 | [6.17, 7.76] | 5.36 | [4.04, 6.67] |
| Period (PS) | 1.9 | [1.40, 2.40] | -0.38 | [-1.06, 0.29] |
| Time (*outtake*) | -6.17 | [-8.57, -3.65] | -4.87 | [-7.53, -2.10] |
| Group (ADHD-W) | -0.25 | [-1.36, 0.85] | 0.88 | [-0.91, 2.68] |
| ROI (parietal) | -0.6 | [-1.10, -0.09] | -0.13 | [-0.80, 0.54] |
| H2 | -0.05 | [-0.23, 0.14] | 0.4 | [0.14, 0.65] |
| Period ×Time | -0.02 | [-0.73, 0.70] | -0.32 | [-1.26, 0.62] |
| Period × Group | -0.47 | [-1.15, 0.23] | -0.51 | [-1.43, 0.40] |
| Time × Group | -0.35 | [-3.68, 2.90] | -0.77 | [-4.33, 2.80] |
| Period × ROI | -0.82 | [-1.53, -0.12] | -0.69 | [-1.63, 0.24] |
| Time × ROI | -0.83 | [-1.54, -0.12] | -0.58 | [-1.52, 0.35] |
| Group × ROI | -0.21 | [-0.90, 0.48] | -0.52 | [-1.44, 0.39] |
| Period × Time × Group | 0.18 | [-0.83, 1.17] | 0.4 | [-0.89, 1.70] |
| Period × Time × ROI | -0.12 | [-1.12, 0.87] | 0.12 | [-1.17, 1.43] |
| Period × Group × ROI | 0.38 | [-0.58, 1.35] | 0.15 | [-1.11, 1.43] |
| Time × Group × ROI | 0.66 | [-0.33, 1.64] | 0.52 | [-0.77, 1.82] |
| Period × Time × Group × ROI | 0.16 | [-1.22, 1.55] | 0.00 | [-1.78, 1.78] |
| ***Note*:** Estimates are posterior means. Reference levels: Period = baseline (BL), Time = *intake*, Group = ADHD-T, ROI = frontal, Half = H1. | | | | |

| **Tabel S5: Convergence diagnostics for Bayesian LMMs.** | | | | | |
| --- | --- | --- | --- | --- | --- |
| **Model** | **Max** $\hat{\boldsymbol{R}}$ | **Min ESS (bulk)** | **Min ESS (tail)** | **Divergences** | **Max treedepth hits** |
| Behavioural LMM | 1.001 | 2849 | 6165 | 0 | 0 |
| Behavioural LMM with NT comparisons | 1.000 | 2090 | 3389 | 0 | 0 |
| N2 mean amplitude LMM | 1.001 | 3752 | 7914 | 0 | 0 |
| N2 peak latency LMM | 1.001 | 9654 | 17046 | 0 | 0 |
| P3 mean amplitude LMM | 1.001 | 5021 | 9730 | 0 | 0 |
| ERP LMM: P3 peak latency | 1.001 | 6939 | 11797 | 0 | 0 |
| N2 peak latency LMM for sensitivity analysis | 1.001 | 9765 | 17856 | 0 | 0 |
| P3 peak latency LMM for sensitivity analysis | 1.001 | 8293 | 14066 | 0 | 0 |
| N2 mean amplitude LMM for sensitivity analysis | 1.001 | 3861 | 7013 | 0 | 0 |
| P3 mean amplitude LMM for sensitivity analysis | 1.002 | 2946 | 6158 | 0 | 0 |
| ERP mean amplitude LMM with NT comparisons | 1.002 | 1931 | 4465 | 0 | 0 |
| ERP peak latency LMM with NT comparisons | 1.001 | 3694 | 4095 | 0 | 0 |
| Spectral LMM: alpha (8 Hz) | 1.003 | 852 | 1267 | 0 | 0 |
| Spectral LMM: theta (4 Hz) | 1.007 | 604 | 1867 | 0 | 0 |
| Pre-stimulus alpha LMM (8–12 Hz) | 1.001 | 7355 | 13472 | 0 | 0 |
| ***Note*.** All models were fitted in brms using NUTS with 4 chains (12,000 iterations; 3,000 warmup; 36,000 post-warmup draws). *R̂* values were ≤ 1.007. No divergent transitions and no maximum treedepth hits were observed. | | | | | |
